# Multimodal prehabilitation enhances innate antitumor immunity via NK cell recruitment

**DOI:** 10.1101/2024.08.05.24311508

**Authors:** Lixuan Feng, Benjamin Gordon, Xin Su, Ariane Brassard, Iqraa Dhoparee-Doomah, Sabrina Leo, Rashami Awasthi, France Bourdeau, Betty Giannias, Heather Gill, Enrico Minnella, Lorenzo Ferri, Sara Najmeh, Jonathan Spicer, Francesco Carli, Jonathan Cools-Lartigue

**Affiliations:** Department of Microbiology and Immunology, McGill University, Montreal, Quebec, Canada; Department of Thoracic Surgery, McGill University, Montreal, Quebec, Canada; Department of Surgery, McGill University, Montreal, Quebec, Canada; Department of Anesthesia, McGill University, Montreal, Quebec, Canada

**Keywords:** Lung cancer, prehabilitation, immunity, NK cells

## Abstract

**BACKGROUND:** While the clinical benefits of multimodal prehabilitation in cancer patients are well defined, the underlying immune modulations have not been studied. The objective of this study was to examine how prehabilitation can alter lung cancer immunity.

**METHODS:** Newly diagnosed lung cancer patients were referred to the prehabilitation clinic for preoperative personalized multimodal intervention (exercise training, nutritional optimization, and anxiety reduction) and blood samples were collected at baseline and surgery. Tumor samples were collected at surgery and compared to matched control samples from patients who did not receive prehabilitation. An animal model was used to study prehabilitation and tumor growth kinetics.

**RESULTS:** Twenty-eight lung cancer patients who underwent multimodal prehabilitation were included (McGill University Health Centre Research Ethics Board #2023-9005). After prehabilitation, patient-isolated peripheral blood mononuclear cells (PBMCs) showed significantly increased cytotoxicity against cancer cells (*p* < 0.0001) and significantly increased circulating natural killer (NK) cells in cohort (*p* = 0.0290) and paired analyses (*p* = 0.0312). Compared to matched controls, patients who received prehabilitation had significantly more intra-tumor NK cells (*p* = 0.0172). *In vivo*, we observed a significant increase in circulating NK cells (*p* = 0.0364) and slower tumor growth (*p* = 0.0396) with prehabilitation. When NK cells were depleted in prehabilitated mice, we observed a decrease in the protective effects of prehabilitation (*p* = 0.0314) and overall, we observed a significant correlation between circulating NK cells and reduced tumor volume (*p* = 0.0203, r = -0.5143).

**CONCLUSIONS:** Multimodal prehabilitation may play a role in antitumor immunity by increasing peripheral and tumour-infiltrating NK cells leading to a reduced cancer burden. Future studies on the protective effect of prehabilitation on postoperative immunity should be conducted.

## Background

Lung cancer remains the leading cause of cancer mortality in the world, accounting for 18% of all cancer deaths (1). Major surgery is often an important treatment modality for lung cancer patients. However, the accompanying surgical stress can be followed by feelings of fatigue and a decreased quality of life, and one study reported a 40% reduction in functional capacity (2).

Multimodal prehabilitation is a novel method for optimizing functional capacity while waiting for surgery and aims to facilitate the recovery process. It includes exercise training, nutritional optimization, and psychological support. This multidisciplinary intervention has been demonstrated to increase physiological reserve before surgery and allow patients to recover faster (3). Functional capacity has been shown to be an independent prognostic factor of survival in surgically treated non-small cell lung cancer patients and is a common way to evaluate prehabilitation (4). In a meta-analysis of 21 studies of 1564 patients, it was determined that prehabilitation exercise for cancer patients significantly improved the 6 minute walking test (6MWT) distance in patients who had underwent prehabilitation (5). Among lung cancer patients, a personalized, stepped prehabilitation program significantly improved patient 6MWT distance significantly by 29.9m, and the length of stay was 2 days in prehabilitated patients compared with 3 days in the usual care group (6). In another study with 15 patients who were enrolled in either multimodal prehabilitation or control, the researchers found that there was a significant improvement in functional capacity among the patients who underwent prehabilitation (7).

Within multimodal prehabilitation, nutrition is also an important component in addressing cancer patient sarcopenia and malnutrition, which are predictors of postoperative mortality and morbidity (8–10). Additionally, a greater risk of lung cancer risk is associated with a poor diet, suggesting that diet plays an important role within the context of cancer (11).

Psychological support and anti-anxiety strategies are important aspects of multimodal prehabilitation as stress and psychological adversity have been previously associated with both higher lung cancer incidence and worse survival (12). Chronic stress may induce immune dysregulation leading to inflammation which has been correlated with tumor growth, progression, and metastasis (13). Functional capacity appears to be a strong predictor of survival within non-small cell lung cancer patients (14).

The oncological benefits of prehabilitation on lung cancer are less well known. In human studies, there has been inconsistent evidence supporting the modulation of the immune system towards anticancer immunity after exercise (15, 16). In studies on human breast cancer and prostate cancer, there is variability in the number of training sessions and large differences in exercise doses, leading to inconsistent findings. In animal studies, exercise appears to suppress tumor growth through mobilization of NK cells through epinephrine and interleukin-6 (17) but in a highly dose dependent manner (18).

Given the potential benefits of multimodal prehabilitation in modulating the immune system toward anticancer immunity, it is important to study how this program may have oncological benefits in lung cancer patients. Therefore, the purpose of this study was to examine how the immune system may be modulated in lung cancer patients after a multimodal prehabilitation program. We hypothesized that lung cancer patients who undergo a prehabilitation program would have an immune phenotype that shifts toward antitumour immunity.

## Methods

### Patient samples and informed consent

This study was conducted in accordance with the *Tri-Council Policy Statement: Ethical Conduct for Research Involving Humans* (*2014*), as well as with the requirements set out in the applicable standard operation procedures of the Research Institute of the McGill University Health Centre and was approved by the McGill University Health Centre Research Ethics Board (2023–9005). Samples were obtained from patients who had consented to participate in the Thoracic Oncology Clinical Database and Biobank (2014–1119). All patients were adults who were scheduled for lung cancer resection between August 2021 and April 2023, and who did not receive neoadjuvant treatment prior to surgery. Patients with metastatic cancer, insufficient comprehension of French or English, or premorbid conditions that contraindicated exercise (severe cardiovascular or neuromuscular diseases) were excluded.

### Prehabilitation program

Patients in the multimodal prehabilitation program were seen approximately 4 weeks before their scheduled operation, at the initial baseline visit by a certified kinesiologist who screened, assessed, trained, and prescribed a personalized structured exercise program for each participant based on their individual assessment. The home-based training included moderate intensity aerobic training 5 days per week for 30 minutes, resistance exercises performed using elastic bands, and flexibility exercises. The program was continued until the patient underwent surgery.

If indicated, a registered dietitian also conducted a comprehensive dietary assessment and nutrition-focused physical exam. Based on these assessments, an individual plan was devised to meet each patient’s nutritional needs. Instructions on how to eat well-balanced meals with a focus on 1.2-1.5g/kg body weight protein intake were also given. Supplementation was prescribed as needed to adhere to the recommendations of the European Society for Clinical Nutrition and Metabolism (ESPEN) (19).

At baseline, patients also met with psychology-trained personnel (nurses with psychosocial specialization) and were provided with techniques aimed at reducing anxiety. These activities included relaxation exercises and deep breathing exercises. Additional sessions were provided as needed.

### Blood draw

Blood was drawn from prehabilitation patients at baseline, prior to the commencement of the prehabilitation program, and immediately before surgery. Baseline samples were used as controls to compare the changes after the program as patients referred to prehabilitation may not have equivalent functional capacity as patients not indicated for prehabilitation.

### Peripheral blood mononuclear cell (PBMC) isolation

PBMC isolation was performed within 6 hours of blood collection using Histopaque-1077 (Sigma-Aldrich) from blood collected in EDTA tubes. PBMCs were washed and resuspended in sterile Dulbecco’s phosphate-buffered saline solution without calcium and magnesium (WISENT). After isolation, the cells were placed on ice for further analysis.

### PBMC cytotoxicity assay

5 x 10^4^ A549-GFP human lung adenocarcinoma cells were grown in DMEM/F12 media (WISENT) supplemented with 10% FBS and 1% penicillin/streptomycin and plated in a 96-well plate and allowed to adhere overnight. PBMCs isolated from patients (2.5 x 10^5^) were resuspended in A549-GFP growth media and plated in sextuplicate with cancer cells. After 2 days, the plates were imaged using an EVOS M500 Imaging System and images were analyzed using ImageJ software. Cell growth was standardized to that of A549-GFP only wells from each repeat experiment.

### Flow cytometry analysis of human PBMCs

Flow cytometry analysis was performed on live patient isolated PBMCs. Live cells were first stained on ice with eBioscience^TM^ Fixable Viability Dye eFluor^TM^ 780 (ThermoFisher) before being blocked with BD Pharmingen^TM^ Human BD Fc Block to reduce nonspecific antibody binding (BD Biosciences). Cells were then stained for CD3-BV650 (BD Biosciences), CD11b-BUV395 (BD Biosciences), CD14-AF488 (BD Biosciences), CD16-BV510 (BioLegend), CD56-PerCP-Cy5.5 (BD Biosciences), CD66b-APC (Miltenyi Biotec) and HLADR-PE-Cy7 (BioLegend). Samples were acquired in the immunophenotyping platform at the Research Institute of the McGill University Health Centre and analyzed using FlowJo version 10.7.2.

### NK cell cytotoxicity assay

5 x 10^4^ A549-GFP human lung adenocarcinoma cells were grown in DMEM/F12 media (WISENT) supplemented with 10% FBS and 1% penicillin/streptomycin, plated in a 96-well plate and allowed to adhere overnight. NK cells were isolated from patient samples using the EasySep^TM^ Direct Human NK Cell Isolation Kit (Stemcell Technologies) and counted using a hemocytometer. NK cells were then resuspended in A549-GFP growth media and plated in triplicate with cancer cells. After 2 days, plates were imaged using an EVOS M500 Imaging System, and images were analyzed using ImageJ software. Cell growth was standardized to that of A549-GFP only wells from each repeat experiment.

### Imaging mass cytometry analysis

Datasets of cell frequencies were obtained from a previously published study on single-cell spatial landscapes of the lung tumor immune microenvironment (20). Imaging mass cytometry was used to characterize the tumor landscape in 832 samples (416 patients) of which 206 may have received prehabilitation. After identifying samples from patients who received prehabilitation, 6 samples were identified to be used for analysis. Control samples matched for staging and predominant histology were then used for statistical analysis.

### Immunofluorescence of human samples

4 µm sections of formalin-fixed paraffin embedded tumour tissue were used for immunofluorescence analysis and stained with anti-human antibodies against NK cells (NKp46; ThermoFisher) and cancer cells (pan-cytokeratin; ThermoFisher). The tissue sections were deparaffinized in xylene and rehydrated in decreasing concentrations of ethanol. Antigen retrieval was then performed using a citrate buffer solution (pH=6). Tissue sections were then permeabilized using 0.1% Triton X and incubated with blocking buffer before adding the primary antibody for 1 hour at room temperature. Slides were washed and then secondary antibodies were added before Vectorshield (Vector Laboratories) was added. NK cell counts were measured by counting positively stained cells in ten random 20x fields within the tumor. Samples were considered positive for NK cell infiltration if an NK cell could be observed in the ten random fields.

### Mouse treadmill exercise model

Seven- to ten-week-old C57BL/6 male mice were used in the study. These mice were procured from Charles River and bred at the Research Institute McGill University Health Centre Animal Facility. The LLC1 mouse lung cancer cell line was grown in DMEM medium (WISENT) supplemented with 10% FBS, and 1% penicillin/streptomycin at 37deg Celsius in a humidified 5% CO_2_ incubator. A total of 2.5 x 10^5^ cells/mouse in a 100µL solution of PBS were injected subcutaneously into the left flank of the mice on day 0. Mice were then split into control or exercise groups and the exercise protocol started on day 1. Exercised mice were involuntarily placed on a rodent 5-lane treadmill (exercise or depletion; Harvard Apparatus), for 30 minutes per day at a gradual increase to 28cm/s, for 6 days/week. Mice were euthanized and tumor and blood samples were taken on day 25 following cancer injection. Whole blood was lysed using BD Pharm Lyse^TM^ lysing solution and stained first on ice with eBioscience^TM^ Fixable Viability Dye eFluor^TM^ 780 (ThermoFisher) for live cells before staining for CD3-PE-Cy7 (ThermoFisher), CD11b-BV510 (BioLegend), Ly6C-PerCP-Cy5.5 (ThermoFisher), Ly6G-AF488 (BioLegend), CD43-BUV395 (BD Biosciences), and NK1.1-PE (Biolegend). Samples were acquired in the immunophenotyping platform at the Research Institute of the McGill University Health Centre and analyzed using Flowjo version 10.7.2.

Where indicated, neutralizing antibodies directed against NK cells (3µg/dose, anti-NK1.1, BioXcell) or saline vehicle control were administered intraperitoneally starting 1 day prior to cancer cell injection and continuing every 3 days. All mice were provided with normal drinking water and had free access to a standard diet. Animal protocols and procedures were approved by the McGill University Health Centre Animal Research Division (Animal Usage Protocol: MUHC-8210).

### Statistical analysis

All data was analyzed using GraphPad Prism 9.2 (GraphPad Software, La Jolla, CA). Data are presented as individual values with standard error of the mean unless otherwise indicated. Significance was determined either using a student’s *t*-test or a Welch’s *t*-test where indicated. ANOVA was used for multi-variate comparisons with Sidak’s multiple comparison done *post hoc* unless otherwise indicated. Throughout, a *p* value of 0.05 was considered statistically significant.

## Results

### Prehabilitation cohort and adherence to prehabilitation

A total of 28 patients were referred to the Montreal General Prehabilitation Clinic between August 2021 and April 2023 and were enrolled in the study. The baseline demographic and clinical characteristics of the patients included within the study are presented in **Table 1**.

**Table 1.** Demographic, Clinical, and Operative Data.

| Variable | Entire Cohort (48) | Control (20) | Prehabilitation (28) | *P* value |
| --- | --- | --- | --- | --- |
| **Age, mean (SD)** | 70 (8.2) | 70 (9.7) | 70 (6.9) | 0.3161<br>(t-test) |
| **Male** | 18 (37.5) | 8 (40.0) | 10 (35.7) | 0.7723 |
| **Histology** |  |  |  | 0.0592 |
| Adenocarcinoma | 31 (64.6) | 17 (85.0) | 16 (57.1) |  |
| Other | 17 (35.4) | 3 (15.0) | 12 (42.9) |  |
| **Pathological Staging** |  |  |  | 0.0837 |
| I | 18 (37.5) | 6 (30.0) | 13 (46.4) |  |
| II + III | 19 (39.6) | 12 (60.0) | 8 (28.6) |  |
| Unknown | 11 (22.9) | 2 (10.0) | 7 (25.0) |  |
| **Surgery Type** |  |  |  | 0.5553 |
| Lobectomy | 32 (66.7) | 14 (70.0) | 17 (60.7) |  |
| Other | 16 (33.3) | 6 (30.0) | 11 (39.3) |  |
| **Length of Stay (days)** | 3 (1-4) | 2 (1-4.75) | 3 (1.25-4) | 0.7076<br>(Mann<br>Whitney<br>test) |

Values are n (%) or median (interquartile range) unless otherwise indicated. Unpaired t tests or Mann Whitney tests (indicated) and either Fisher's exact test or chi-square tests were used to calculate *p* values.

The control patients were a matched cohort of historical patient samples. No significant differences were found between the two groups. All patients underwent at least 4 weeks of prehabilitation and did not receive neoadjuvant treatment. Adenocarcinoma was the predominant subtype of lung cancer within our cohort. Most patients underwent lobectomy, and the median length of stay after surgery being 3 days. No adverse events related to prehabilitation were observed. Unless otherwise indicated, within our study control patient samples consisted of baseline blood data from patients who were referred for prehabilitation.

### Prehabilitation enhances PBMC cytotoxicity

As there could be functional capacity differences between patients referred to the prehabilitation program and the historical cohort, we assessed the direct effects of prehabilitation on PBMC cytotoxicity by comparing blood drawn at baseline, prior to the start of the program, to blood drawn immediately prior to surgery. To assess differences in PBMCs after prehabilitation, we first isolated patient PBMCs and plated them with A549-GFP cells at a 50:1 effector-target ratio. After 2 days, the wells were imaged using the EVOS M500 microscope and the percentage of A549-GFP growth was calculated (**Fig. 1**).

**Figure 1.**
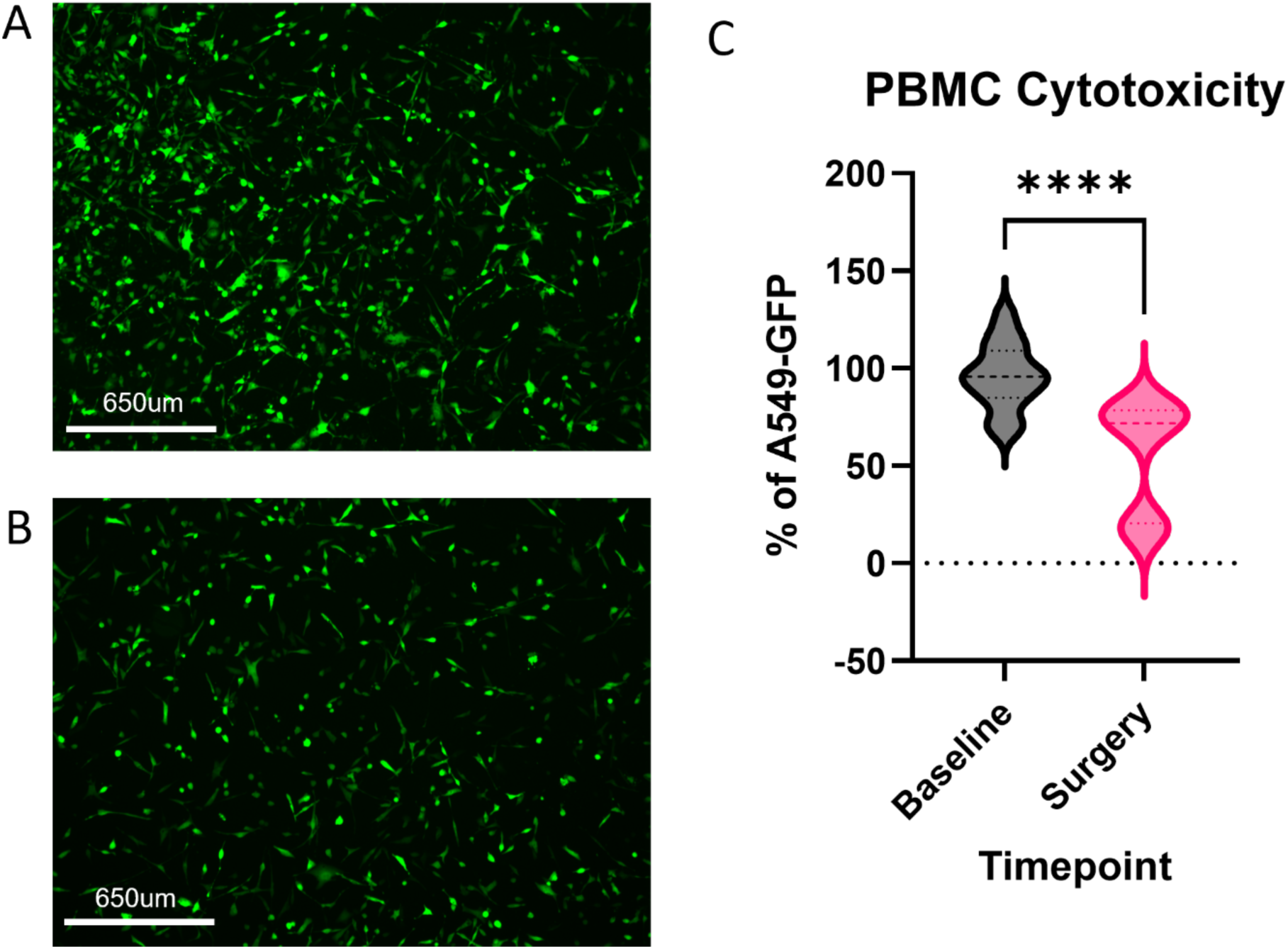
Prehabilitation enhances patient PBMC cytotoxicity. Patient isolated PBMCs were incubated with A549-GFP cells and imaged after 2 days. Growth was calculated with ImageJ using a reference A549-GFP only well as reference. All samples were plated in sextuplicate. **A.** Baseline patient PBMCs (n=3). **B.** Surgery patient PBMCs (n=3). **C.** Quantification of remaining percentage of A549-GFP cells standardized to control after 2 days (*p* < 0.0001, Mann-Whitney two-tailed test).

Baseline samples refer to samples taken prior to the commencement of the prehabilitation program while the surgery timepoint refers to samples taken after the program. Compared with baseline control samples, patient samples after prehabilitation at the time of surgery appeared to have increased cytotoxicity (*p* < 0.0001) against A549-GFP cells compared to baseline control samples. PBMCs from patients at baseline prior appeared to be cytotoxic but to a lesser extent than those from patients after prehabilitation. This result suggested that prehabilitation can modulate the PBMC population toward stronger antitumor immunity.

### Prehabilitation increases the percentage of circulating NK cells

We next sought to determine whether the increase in PBMC cytotoxicity observed after prehabilitation was due to the expansion of specific cell populations. Exercise plays a large role in prehabilitation and has been shown to modulate proportions of immune cells within cancer. Therefore, we wanted to determine which cell populations could contribute to our observed results. We used flow cytometric analysis to determine cell population changes of innate immune cells in the PBMC fraction of whole blood. PBMCs were isolated on the day of blood draw and stained for markers for NK cells (CD3^-^CD14^-^CD56^+^CD66b^-^); classical (HLADR^+^CD3^-^ CD11b^+^CD14^+^CD16^-or^ ^lo^), intermediate (HLADR^+^CD3^-^CD11b^+^CD14^+^CD16^+^), and nonclassical monocytes (HLADR^+^CD3^-^CD11b^+^CD14^lo^CD16^+^); mononuclear MDSCs (HLADR^-^CD3^-^ CD11b^+^CD14^+^CD66b^-or^ ^lo^), and polymorphonuclear MDSCs (HLADR^-^CD3^-^CD11b^+^CD14^-^ CD66b^+^).

We found that after prehabilitation, the percentage of circulating NK cells significantly increased (*p* = 0.0161) (**Fig. 2A**).

**Figure 2.**
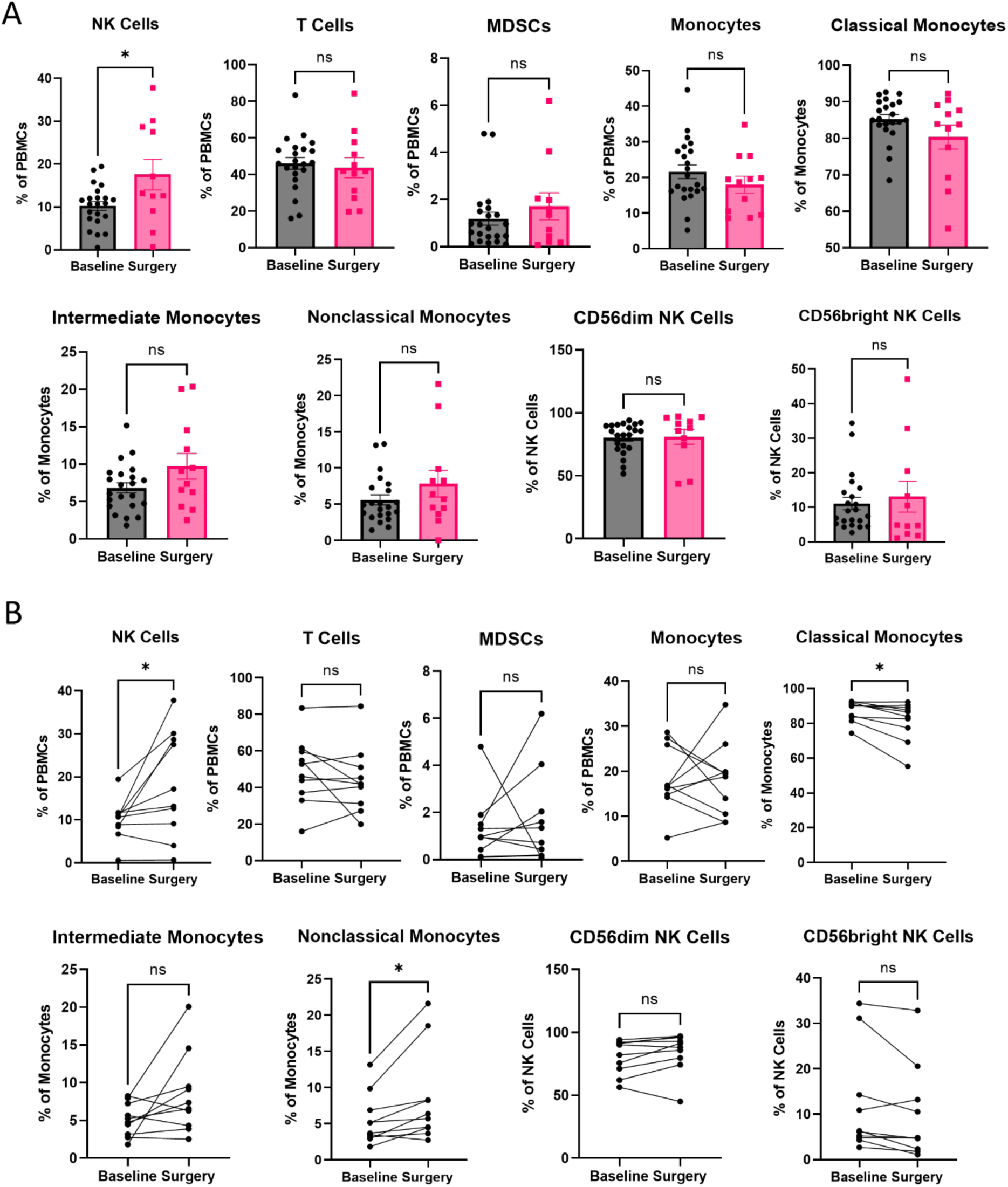
Prehabilitation increases peripheral NK cells and nonclassical monocytes, and decreases classical monocytes. Patient isolated PBMCs were stained for various immune cells and a percentage of total PBMCs was calculated using flow cytometry analysis. **A.** In the cohort analysis, after prehabilitation, the percentage of NK cells of total PBMCs increased significantly (*p* = 0.0161, unpaired t-test). **B.** Within the paired analysis, after prehabilitation the percentage of NK cells increased significantly (*p* = 0.0312, paired t-test) and within the monocyte population, classical monocytes decreased significantly (*p* = 0.0021, paired t-test) and nonclassical monocytes increased significantly (*p* = 0.0188, paired t-test).

As different subtypes of NK cells may play various effector roles such as with CD56dim NK cells being canonically more cytotoxic and CD56bright NK cells playing a larger role in cytokine production, we wanted to determine whether the NK cell fraction changed after prehabilitation. We observed that there were no differences in the percentages of circulating CD56dim or CD56bright NK cells among the total NK cell population. There were no significant differences in the overall percentage of CD3+ T cells or with MDSC population. With the monocyte fraction, we did not observe changes in the total percentage of circulating monocytes or in the subtypes of monocyte within the total monocyte compartment.

As we observed large heterogeneity in the baseline values within our cohort, we also then analyzed ten paired samples from this cohort for which we matched the baseline and surgical timepoint data. Similarly, to the observations within the larger cohort, we observed a significant increase in circulating NK cells (*p* = 0.0312) (**Fig. 2B**) but no differences in the subtype of NK cells. Additionally, we did not observe any differences in the percentage of circulating T cells, MDSCs, or monocytes. However, within the monocyte fraction, we noticed that in our matched samples that there was a significant decrease in the percentage of classical monocytes (*p* = 0.0177) and a significant increase in the percentage of nonclassical monocytes (*p* = 0.0214).

These results indicate that prehabilitation modulates the immune profile of patients who undergo the program by increasing the percentage of circulating NK cells, increasing the percentage of nonclassical monocytes, and decreasing the percentage classical monocytes. Our results suggest that the increase in NK cells may play a role in the increased PBMC cytotoxicity observed. Additionally, nonclassical monocytes are thought to be antitumor and classical monocytes are believed to be protumor; therefore, these findings suggest that prehabilitation modulates the peripheral immune system toward an antitumor protective phenotype.

### Decreasing NK cells leads to decreased PBMC cytotoxicity

As we saw significant differences in NK cells after prehabilitation, we wanted to assess their functionality. NK cells can be cytotoxic against cancer cells, and so we next investigated whether the increased cancer cell killing observed in PBMCs after prehabilitation was due to an increase in NK cells or due to increased NK cell cytotoxicity. NK cells were isolated from fresh patient blood using the EasySep^TM^ Direct NK Cell Isolation Kit (Stemcell Technologies) and plated at various effector-to-target ratios against A549-GFP cells. After 2 days, wells were imaged, and the percentage of A549-GFP growth was standardized to a control.

We observed that as the effector-target ratio of NK cells increased, the killing of A549-GFP cells significantly increased (*p* = <0.0001) (**Fig. 3**).

**Figure 3.**
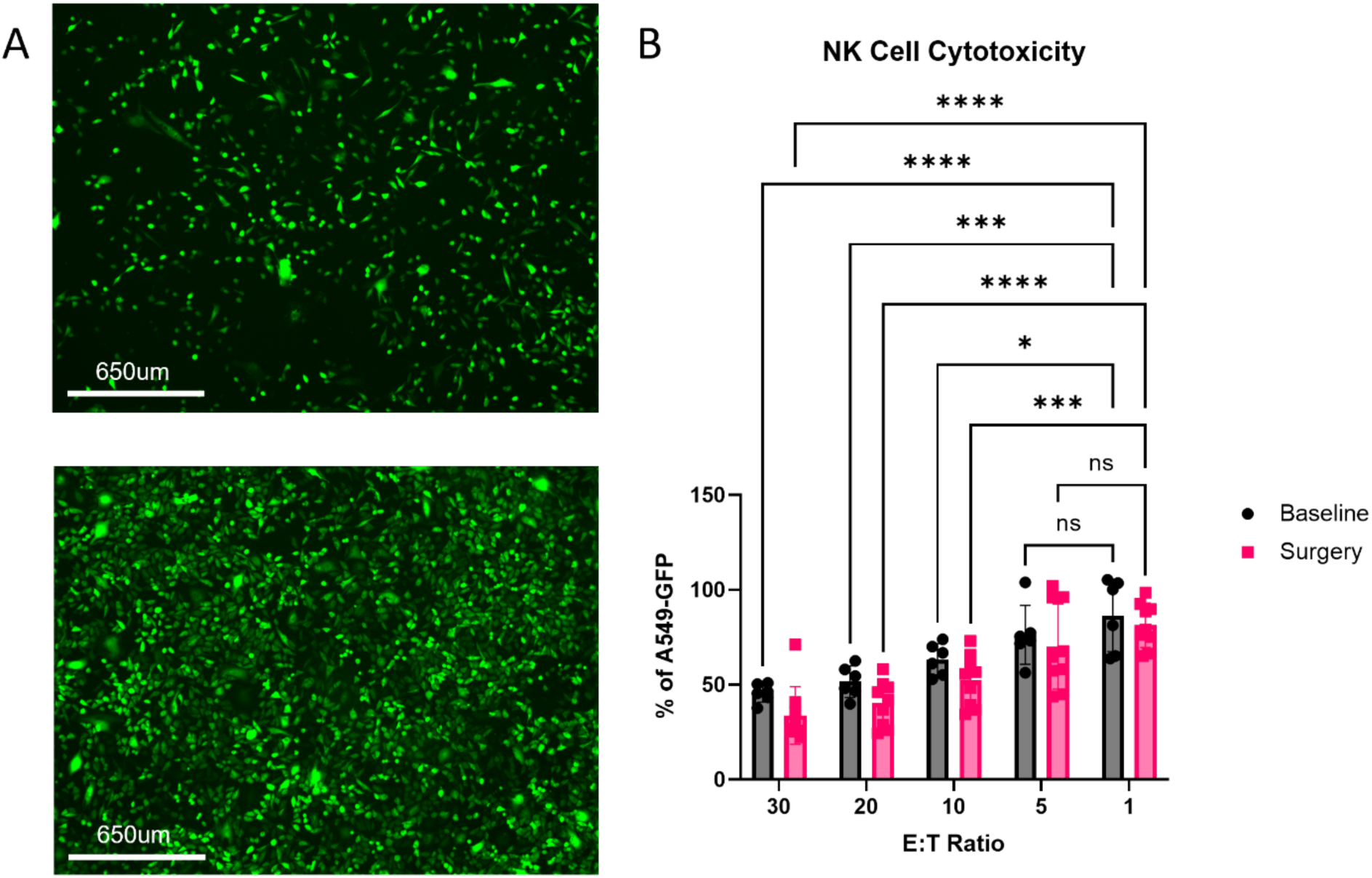
Decreasing amounts of NK cells lead to decreased PBMC cytotoxicity. Patient NK cells were isolated and their cytotoxicity against A549-GFP cells was tested at various effector-to-target ratios (n=2 baseline, n=3 surgery). All samples were plated in triplicate. **A.** Representative image of the 30:1 effector-target ratio. **B.** Representative image of the 1:1 effector-target ratio. **C.** Quantification of remaining % A549-GFP cells standardized to control after 2 days. (*p* = <0.0001, 2way ANOVA followed by Sidak’s multiple comparison test).

However, at the same effector-to-target ratio, there appeared to be no difference between control and prehabilitated patient NK cells. This result suggested that the NK cell-mediated cytotoxicity was due to increased NK cell counts, in contrast to the increased cytotoxic function of individual NK cells.

### Prehabilitation increases NK cell infiltration into tumor tissue

To determine whether the increase in peripheral NK cells had an impact on the primary tumor, we used data from a previously published study on the tumor immune microenvironment in lung cancer (20). We identified patients who received prehabilitation from these historical datasets. Of the 206 samples, 6 samples from 3 patients were identified to have received prehabilitation. For our control cohort of 18 samples from 9 patients, we matched for staging and predominant histology. Given the limited sample size, we did not match for age. We found that there were no significant differences in the percentage of cancer cells between the prehabilitation group and the control group (**Fig. 4A**).

**Figure 4.**
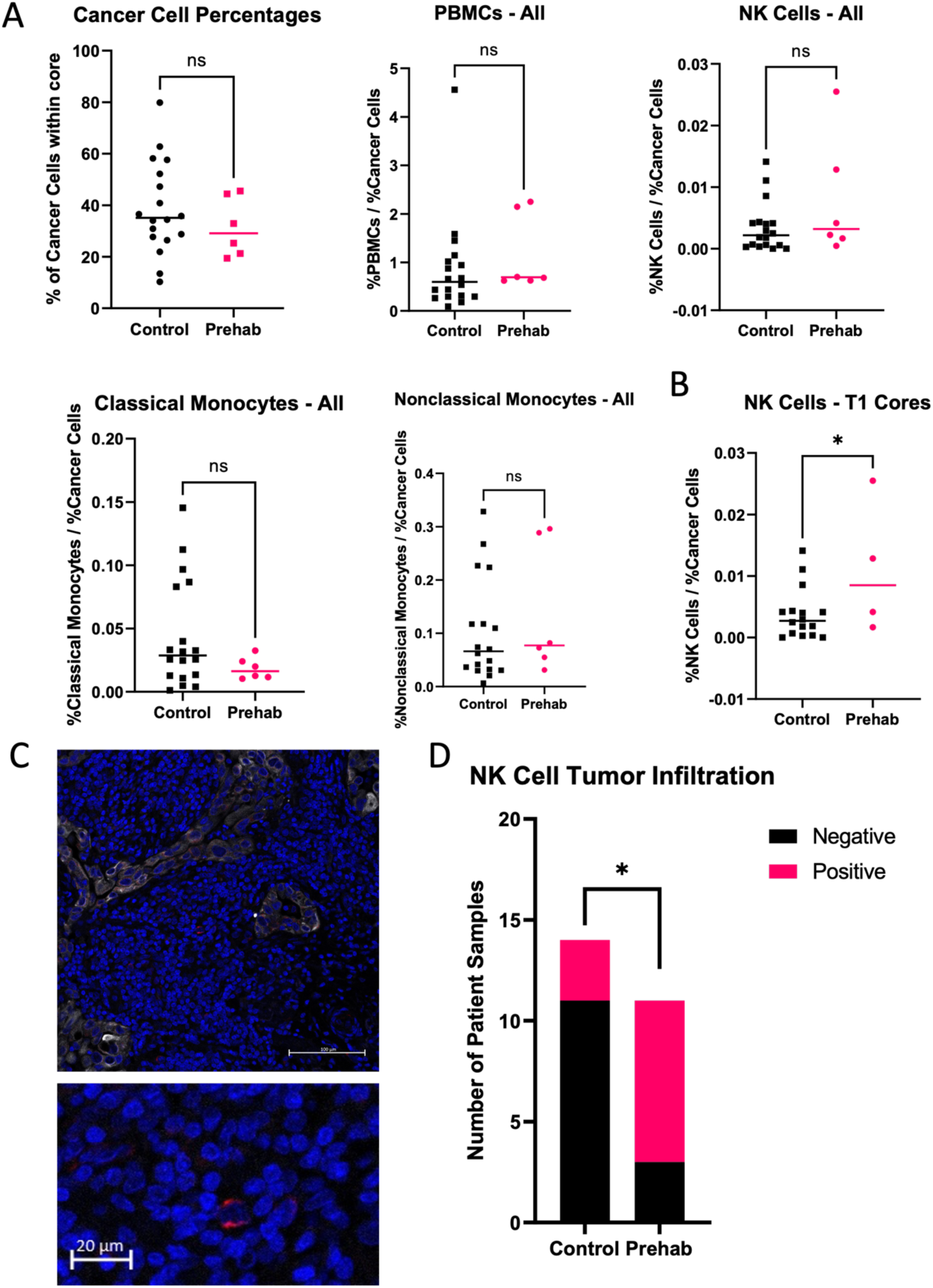
Prehabilitation increases NK cell infiltration into tumor tissue. Using a previously published dataset on lung cancer immune microenvironments, cell percentages were calculated. **A.** Cancer cell and immune cell percentages of samples from patients who underwent prehabilitation (n = 6) were compared (n = 18) (unpaired t-tests). **B.** In samples from patients with early-stage, non-candidates for neoadjuvant therapy, NK cell percentages of control (n = 16) were compared to prehab samples (n = 4) *(p* = 0.0379, unpaired t-test). Patient formalin-fixed paraffin-embedded (FFPE) lung tumor tissue sections were stained by immunofluorescence for NK cells (red), cancer cells (white), and DAPI (blue). **C.** Representative image of a high powered field. **D.** Quantification of samples positive for NK cells in control and prehabilitation patient samples (*p =* 0.0172, Fisher’s exact test).

Likewise, when comparing the percentage of total PBMCs, there were no significant differences between the two groups in overall percentage of infiltrating immune cells, NK cells, classical monocytes, or nonclassical monocytes. However, after separating the historical cohort into those with early-stage disease who were not candidates for neoadjuvant therapy, consistent with those who received prehabilitation within our study, we found there was a significant increase in the percentage of NK cells found within the tumor immune microenvironment in samples from patients who had received prehabilitation (*p =* 0.0379) (**Fig. 4B**).

To confirm our findings of increased NK cells within the tumor immune microenvironment, we investigated the infiltration of NK cells within our patient cohort. We stained resected patient lung samples from individuals who underwent prehabilitation and compared them to historical control patient samples from individuals who were not referred to and did not undergo the prehabilitation program. We used immunofluorescence and stained for NK cells (NKp46, red), cancer cells (pan-cytokeratin, white), and DAPI (cell nucleus, blue) to quantify the number of NK cells within the lung tissue sections (**Fig. 4C**). Using 10 random high magnification fields from each sample, samples were deemed as positive if an NK cell was found.

Tumor samples from patients who underwent prehabilitation had significantly more NK cell infiltration than did those from historically matched controls (*p* = 0.0172) (**Fig. 4D**). Overall, the number of NK cells observed in our samples was low. This finding suggested that the increase in circulating NK cells observed after prehabilitation could contribute to the increase in the number of tumor-infiltrating NK cells.

### Exercise protects against the loss of NK cells seen with cancer progression

Lung biopsies are not standard within lung cancer care making it difficult to evaluate how prehabilitation may modulate the tumor. Thus, to investigate the direct effect of prehabilitation tumors, we used an *in vivo* mouse model to study tumor growth dynamics. Wild-type mice were injected subcutaneously with LLC1 lung cancer cells and then subjected to a moderate-intensity treadmill-running exercise regimen [6x/week; 30 minutes; gradual increase to 28 cm/s] adapted from previous studies (21, 22) (**Fig. 5A**).

**Figure 5.**
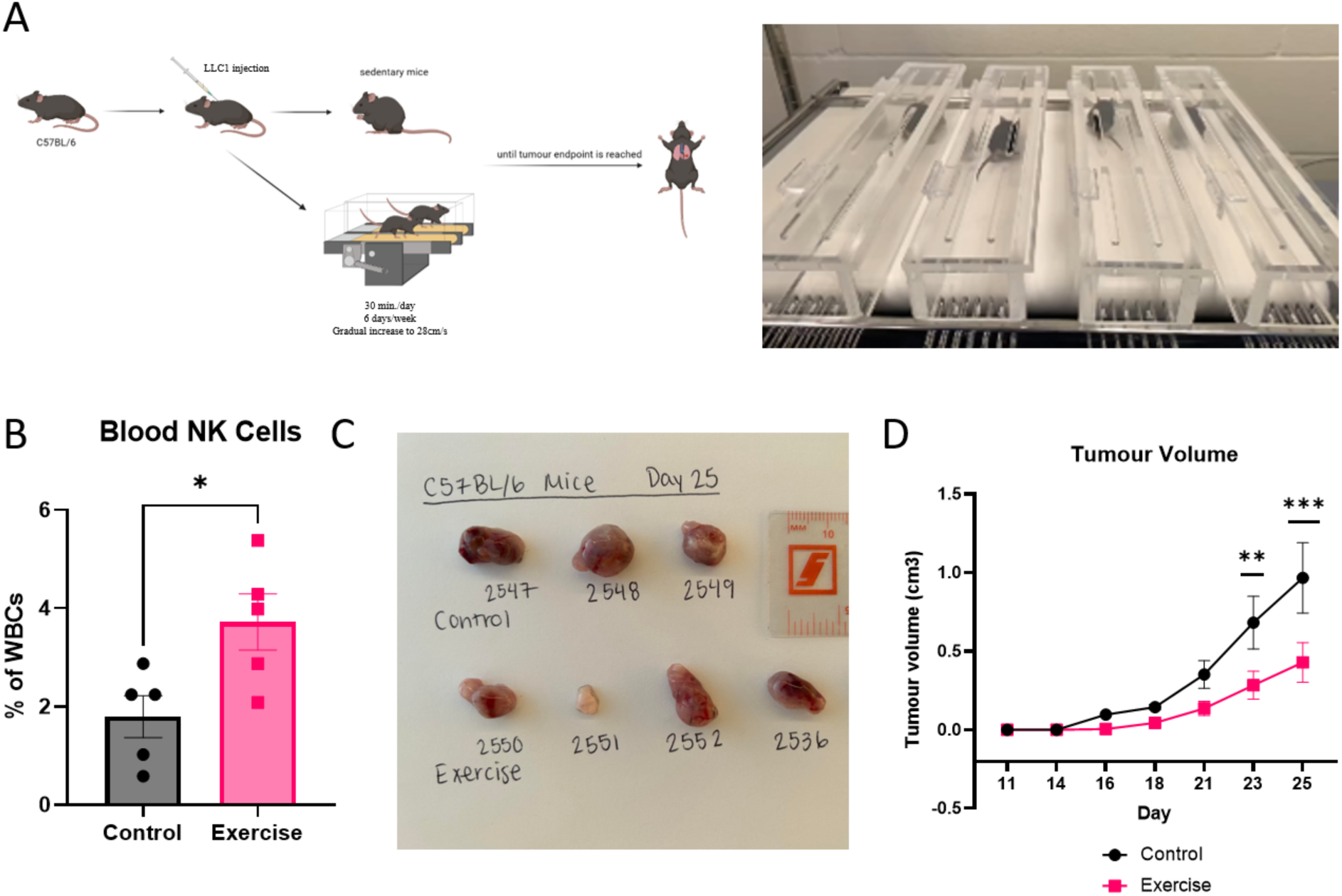
Exercise increases circulating NK cells and reduces tumor growth. A. A schematic of the treadmill running exercise model is shown. C57BL/6 mice were injected subcutaneously in the left flank with LLC1 cells. **B.** Blood taken at endpoint, day 25, shows that exercise has significantly more NK cells (*p* = 0.0270, unpaired t-test). **C.** Representative images of tumor size at endpoint. **D.** Quantification of tumor growth (*p* = 0.0396, 2way ANOVA followed by Sidak’s multiple comparisons).

The prehabilitation program also included nutritional supplementation if indicated for individual patients; thus, all the mice had free access to a standard diet.

Recapitulating our patient data, we observed that within this model, circulating NK cells were significantly increased in our exercised mice (*p =* 0.0270) (**Fig. 5B**). Within our exercised mouse group, exercise also appeared to significantly reduce tumour progression and delay oncogenesis. The control mice had significantly larger tumors then did the exercised mice group (*p* = 0.0396). In addition, the control mice appeared to have more visible tumours earlier than did the exercised mice. These results suggest that exercise may play a role in antitumor immunity by increasing the number of circulating NK cells and reducing the tumor burden. Overall, these findings suggest that the exercise portion of prehabilitation could provide tumor-protective benefits for lung cancer progression.

### NK cells are responsible for exercise-induced delayed tumor progression

Exercise has been demonstrated to play a role not only in NK cell recruitment, but also in various other functions. Therefore, we wanted to assess whether our tumor-protective benefits of exercise were due to increased NK cells. To determine whether the increase in circulating NK cells resulted in delayed oncogenesis and tumor progression, we then depleted circulating NK cells intraperitonially using an anti-NK1.1 antibody (**Fig. 6A**) 1 day prior to cancer cell injection and then every 3 days thereafter. Control and exercise-only mice were injected with a saline vehicle control. The exercise-only and NK cell-depleted mice were then subjected to exercise on a treadmill using the previously established exercise protocol until they were euthanized.

**Figure 6.**
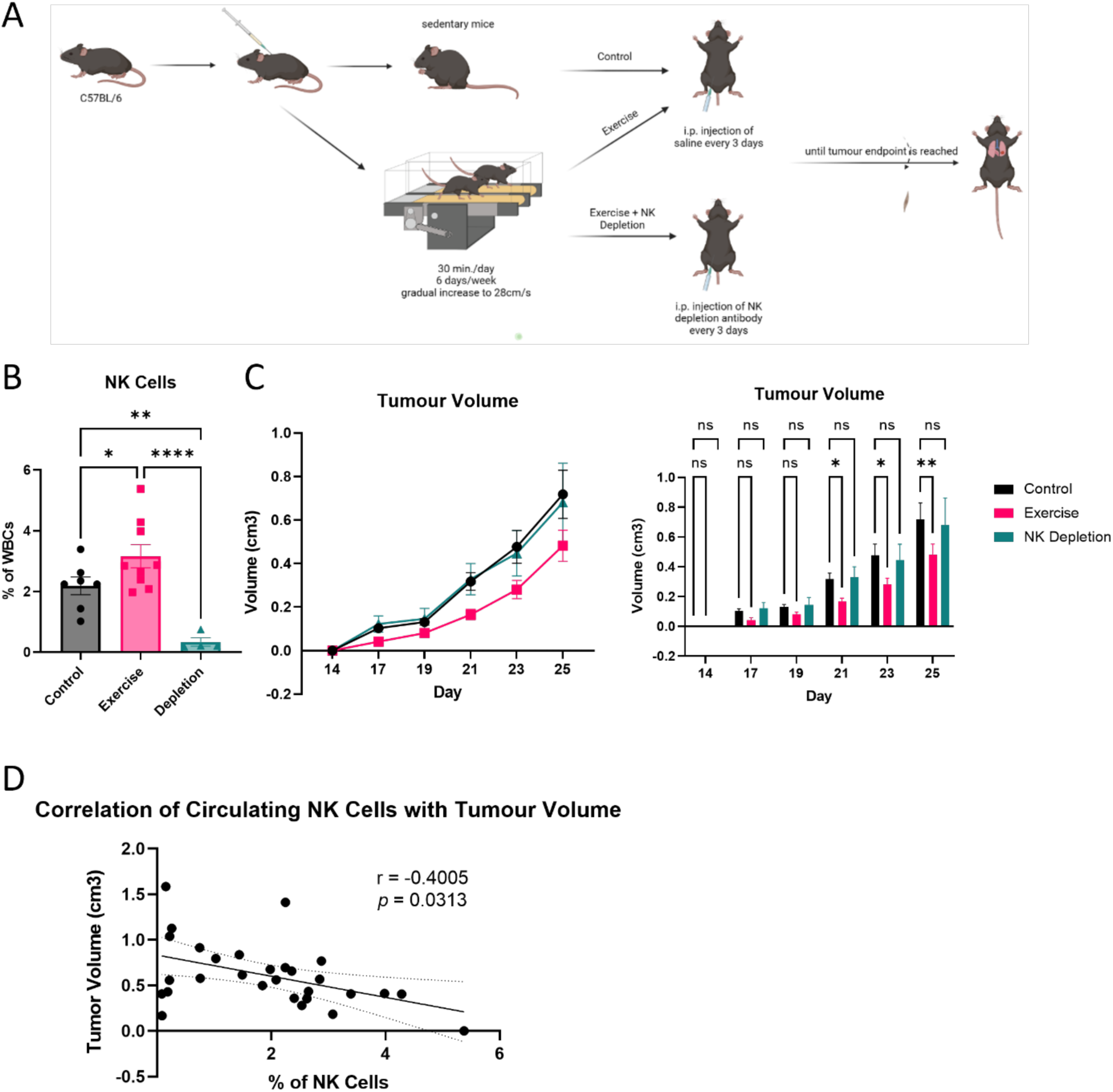
NK cell depletion reduces the antitumor benefits of exercise. **A.** A schematic of the experiment is shown. C57BL/6 mice were injected subcutaneously in the left flank with LLC1 cells and every 3 days were injected either with saline or with a NK depletion antibody until euthanized. **B.** Blood taken at endpoint, day 25, shows that exercise has significantly more NK cells (*p* = 0.0003, one-way ANOVA followed by a Fisher’s LSD test). **C.** Quantification of tumour growth (two-way ANOVA followed by a Fisher’s LSD test). **D.** Spearman’s rank correlation of circulating NK cells with tumour volume is shown (*p =* 0.0313, *r* = -0.4005).

Consistent with our findings from the previous experiment, at the endpoint, we observed significantly fewer circulating NK cells in our control group than in the exercised group (*p =* 0.0003) (**Fig. 6B**). Additionally, the anti-NK1.1 antibody appeared to work to deplete NK cells throughout the experiment.

When assessing tumour growth, we found that exercise led to slower tumour growth than the control (**Fig. 6C**). However, after circulating NK cells were depleted, the mice that were then subjected to exercise no longer exhibited any differences from the control mice. This finding suggested that the reduction in tumor growth observed in he exercised group was mediated by NK cells.

As our results have indicated that exercised mice had increased circulating NK cells and reduced tumor burden, we performed a Spearman’s rank correlation analysis to determine whether the exercise-induced slower tumour progression was linked to circulating NK cells. We found there was a significant correlation between peripheral NK cells and reduced tumor volume suggesting that increased NK cells were correlated with reduced tumor size (*p* = 0.0313, r = - 0.4005) (**Fig. 6D**). Overall, these findings suggest that the exercise-mediated antitumor effects are due to NK cell involvement.

## Discussion

This study revealed a previously undescribed role for exercise-based multimodal prehabilitation in inducing an antitumor immune response through NK cell involvement in lung cancer patients. Multimodal prehabilitation increases *ex vivo* PBMC cytotoxicity which we attributed to increasing circulating NK cells within the total PBMC population. When we studied the cytotoxicity of these NK cells, we found that an increasing effector-to-target ratio led to an increase in NK cell cytotoxicity but that there was no difference in the functional capacity between baseline and after prehabilitation suggesting that the increase in cytotoxicity was directly linked to an increase in NK cells. Looking at historic cohorts from a tumor immune microenvironment dataset, we compared those with early-stage non-candidates for immunotherapy who had received prehabilitation with matched controls and also observed increased NK cells within the tumor. When we compared our prehabilitation patient cohort samples to those of matched controls, we also found significantly increased intratumor NK cell infiltration. Thus, our findings of increased numbers of circulating and intratumor NK cells are especially important and beneficial because these are a subset of immune cells that are known to be impaired and reduced in lung cancer. Although validation in a larger dataset is needed, our data provide some of the first evidence showing that prehabilitation can increase circulating and tumour-infiltrating NK cells and have a direct effect on human lung cancer.

Traditional perioperative interventions are performed in the postoperative period, but recent advances suggest that the preoperative period may also present an important timeframe for improving factors that contribute to enhanced patient recovery. Prehabilitation aims to provide interventions to patients prior to surgery with the aim of optimizing postoperative outcomes (23). The belief is that by enhancing an individual’s functional capacity, this may enable them to better withstand an incoming stressor (24). Functional capacity appears to be a strong predictor of survival within non-small cell lung cancer patients (14). Within lung cancer patients, a personalized, stepped prehabilitation program was found to have improved patient six-minute walking distance significantly by 29.9m and the length of stay was 2 days in prehabilitated patients opposed to 3 days in the usual care group (6). In another study with 152 patients either enrolled in multimodal prehabilitation or control, the researchers found there was a significant improvement in functional capacity within the patients who underwent prehabilitation (7).

Animal models support prehabilitation and increased functional capacity prior to surgery. In a group of rats, mortality was significantly decreased when the rats were previously exposed to exercise prior to a trauma and the animals showed increased resistance to trauma (25). In mouse studies, prehabilitative exercise was successful in ameliorating cancer cachexia, improving survival, and in reducing tumor growth in mice (26, 27). With preoperative exercise therapy, one systemic review determined that preoperative exercise therapy was effective in reducing postoperative complications while another study demonstrated that prehabilitation improved postoperative pain and physical function (28, 29). Multimodal prehabilitation including exercise training, nutritional support, and psychological support which has been demonstrated to increase physiological reserve before surgery and appears to allow patients to have a faster recovery (3). Exercise plays a large role in multimodal prehabilitation as it is implemented for all patients whereas nutritional supplementation and psychological support are given if indicated.

As exercise has been previously demonstrated to suppress tumor growth through epinephrine- and IL-6-dependent NK cell mobilization, we also studied the exercise component of prehabilitation within a lung cancer animal model to determine how exercise may change tumor growth kinetics (17). The moderate exercise protocol in this study was chosen in consideration of the functional status of patients referred to the prehabilitation clinic. When evaluated *in vivo* using a forced treadmill exercise mouse model, we were able to recapitulate our findings of increased circulating NK cells after exercise. Additionally, we observed a slower progression of tumor growth and a significant correlation between circulating NK cells and reduced tumor size suggesting that increasing circulating NK cells can be a cancer protective effect of exercise.

Our study focused mainly on how prehabilitation influences NK cells, but recent studies in prostate cancer (16), pancreatic cancer (22), and breast cancer (15) have suggested that the type of cancer also plays a role in which immune cell populations are modulated with exercise. Other immune cell types such as CD8+ T cells or FOXP3+ Tregs may also play a role and would be important to be studied in future projects. Within our prehabilitation cohort, no patients received neoadjuvant chemo- or immune-therapy. However, given the extensive research into the use of these modalities prior to surgery, it would be important for future studies to evaluate how prehabilitation may work in conjunction with neoadjuvant therapies. Given that several studies have determined that prehabilitation can modulate the immune system, our findings corroborating this, the potential role for prehabilitation to sensitize the tumour-immune environment to these treatments is not to be understated and an important future avenue to study.

Due to the scope of this project, we focused our study on NK cells. However, within our paired patient samples, we did also observe that classical monocytes significantly decreased while nonclassical monocytes significantly increased. This could be an important future avenue to explore, as classical monocytes are thought to migrate from the blood into the tissue to polarize into protumor TAMs, suppress T-cell function, and remodel the extracellular matrix while nonclassical monocytes may engulf tumor material and inhibit Tregs (30–34).

The present study has several limitations. Our data are based on a small sample size, and thus, the results are prone to type 2 errors. Within the mass cytometry dataset, the sample numbers were small, as it was not feasible within the timeframe of the current study to investigate our patient samples; therefore we looked at historical controls instead. To our knowledge, this data analysis is the only one that exists and so was included. Additionally, multimodal prehabilitation is a nonstandardized modality that is personalized to patients As such, variations in nutritional supplementation, aerobic and anaerobic exercise, and psychological status could all lead to heterogeneity within our findings, which may lead to our results not being directly applicable to all lung cancer patients. This study also focused heavily on the exercise component of multimodal prehabilitation but equally as important to study are the nutrition and psychological support components.

Nutrition plays a large role in the microbiome of the host and can play a significant role within the context of cancer. Altered microbiome configurations from poor nutrition can lead to dysbiosis and influence cancer promotion through direction interaction of individual microbes (35) or through secreted or modulated metabolites (36, 37). Beyond the microbiome, diet can promote inflammation and drive dysregulated immune responses and poor dietary patterns have been associated with higher lung cancer risk. Likewise, psychological adversity and stress are associated with higher lung cancer incidence and worse survival (12). One potential explanation is that psychosocial factors might promote high-risk behaviours such as smoking, poor diet, obesity, excessive alcohol consumption, and poor sleep. Another proposed explanation is that stress can induce immune dysregulation and thus lead to alterations in inflammation and thereby tumour growth, progression, and metastasis (13). While one limitation of our study is that we did not address the effects of the individual modalities that comprise prehabilitation and within our *in vivo* experiments mainly focused on exercise, likely multimodal prehabilitation has beneficial implications for patients beyond solely the benefits driven from exercise as nutritional and psychological support have important implications for cancer.

## Conclusions

In summary, 28 lung cancer patients underwent multimodal prehabilitation prior to surgery. Prehabilitation patients showed significantly increased PBMC cytotoxicity, increased circulating NK cells, and increased tumor-infiltrating NK cells. When the effects of prehabilitation on tumour growth kinetics were studied *in vivo*, we observed a similar increase in circulating NK cells and reduced tumour growth. The antitumor effects appeared to be NK cell driven, as the protective benefits were no longer observed when NK cells were depleted in exercised mice. This study provides the first evidence in humans that multimodal prehabilitation can directly affect human lung cancer, providing insight into the immune modulations through which prehabilitation can promote antitumor immunity in lung cancer patients. While future work will be required to determine the feasibility of prehabilitation for patients with lung cancer on a larger scale, our findings demonstrate that prehabilitation can promote antitumor immunity and can lead to new approaches for the treatment lung cancer.

## List of abbreviations

6MWD: 6-minute walk distance (m)
6MWT: 6-minute walk test
A549: human lung adenocarcinoma cell line
ECM: extracellular matrix
FFPE: formalin-fixed paraffin-embedded
GFP: green fluorescent protein
HIIT: high-intensity interval training
LLC1: Lewis Lung cancer mouse cell line
MDSC: myeloid derived suppressor cell
NK: cell natural killer cell
NSCLC: non-small cell lung cancer
PBMC: peripheral blood mononuclear cells
TAM: tumour associated macrophage
TME: tumour microenvironment

## Declarations

### Ethics approval and consent to participate

This study was conducted in accordarce with the *Tri-Council Policy Statement: Ethical Conduct for Research Involving Humans* (*2014*), as well as with the requirements set out in the applicable standard operation procedures of the Research Institute of the McGill University Health Centre and was approved by the McGill University Health Centre Research Ethics Board (2023–9005). Samples were obtained from patients who had consented to participate in the Thoracic Oncology Clinical Database and Biobank (2014–1119). All patients were adults who were scheduled for lung cancer resection between August 2021 and April 2023, and who did not receive neoadjuvant treatment prior to surgery.

### Consent for publication

Consent was obtained directly from patient(s) as set out in the Thoracic Oncology Clinical Database and Biobank consent forms (2014–1119).

### Availability of data and material

The datasets generated and/or analyzed during the current study are available from the corresponding author on reasonable request.

## Acknowledgements

We would like to acknowledge and thank the Spicer lab for allowing us to use their lung cancer dataset for analysis.

## Authors’ contributions

LF and JCL designed the study. FL, RA, and FC recruited the patients. LF, BG, XS, AB, IDD, SL and investigators collected data. LF and JCL developed the statistical analysis plan. LF, FC, and JCL wrote the draft of the manuscript. All authors provided final approval to submit the manuscript for publication.

